# Prevalence of hepatitis B and hepatitis C in migrants from sub-Saharan Africa at Al Kufra in SE Libya in transit toward Europe

**DOI:** 10.1101/2022.02.11.22270214

**Authors:** Frhat M. A. Saaed, Jerry E. Ongerth

## Abstract

**Background:** Viral hepatitis has become a universally established health care challenge due to its worldwide distribution chronic persistence, complications, and high prevalence with unchecked conditions in areas like sub-Saharan Africa. A high proportion of asymptomatic infections allows serious complications in patients who are infected. These asymptomatic infected people pose a serious risk for the transmission of infection to healthy population. This study aimed to detect and examine the prevalence of both HBV and HCV carriers among 3248 newly arrived migrants from different parts of sub-Saharan Africa in Al Kufra, Libya, a remote agricultural North African city. All these migrants were required by the Libyan authorities to undergo a complete medical check-up for different purposes such as joining for new jobs, and for obtaining licenses for trade and commerce.

**Methodology:** UAT sera from 3,248 migrants, aged 18–53 years, attending the Al Kufra city hospital from January 01 to December 31, 2019, were screened for HBsAg and anti-HCV antibody by rapid tests and positive samples were further tested by ELISA method.

**Results:** **t**he results showed that 761/3248 (23.4%) of the migrants were positive for HBV and 1014/3248 (31.2%) were positive for HCV.

**Conclusion:** Migrants newly arrived from sub-Saharan African carry high rates of Hepatitis B and C infections. It is important to increase awareness about occupational health, and the risks of HBV and HCV transmission to the local population. The study results indicate the value of preemployment medical check-up and regular investigation and illustrates the importance of understanding the potential impact on migrant destination populations.

## Introduction

Hepatitis is a worldwide health problem having important geographic distribution features affecting selected populations disproportionately with resulting implications associated with patterns of migration from economically disadvantaged and security threatened regions. Of particular importance are the blood- and semen-borne hepatitis infections, hepatitis B (HBV) and hepatitis C (HCV). World Health Organization [32] estimates worldwide nearly 300 million HBV infections accounting for nearly 800,000 deaths, and about 60 million HCV infections accounting for 300,000 deaths. The highest endemic HBV and HCV concentrations are in tropical and subtropical regions of Africa, Asia, and the Americas [6]. Combined with social and economic migration pressures from Africa and the Middle East, attention has been focused on the flow of infectious diseases in migrant populations. Libya, in its geographic position between sub-Saharan Africa and Europe, is a focal point for migrating populations from high endemic HBV and HBC regions to the South. This study sought to quantify the extent of these infections in current migrant populations at a major migration route transit point toward Europe.

Migrants and refugees from troubled African regions funnel through sub-Saharan areas and travel poorly established routes through trackless desert to reach the farthest Southeastern Libyan city, Al Kufra, and the road leading to the Mediterranean [30, 22]. Al Kufra city is the located in southeast Libya, about 1000 km to the nearest point on the Mediterranean coast, and 1700 km by road from Tripoli, the capital city and shortest route to Italian territory. Libya has international borders with Egypt in the east and with Sudan and Chad in the south. Migration routes to and through Libya are complex and vary to meet changing circumstances as described by the UNHCR [30].

The Al Kufra district, roughly the size of Spain, is the remote Southeastern quarter of Libya. It is at the beginning of the road connecting Al Kufra City to the North. No roads extend farther to the borders of Egypt to the east, the Sudan and Chad to the South, approximately 200, 300, and 400 km away respectively, across open roadless desert, Figure 1. The majority of migrants from different parts of Africa arriving at Al Kufra from illegal border crossings, have no required documents or work permits. Highest migrant origins include Sudan and Chad (Central Africa), the next greatest proportion of migrants entering the city originated from Burkina Faso, Niger, Mali, Nigeria, and Ghana (West Africa), and from Eretria, Somalia, and Ethiopia (Horn of Africa). Departing from the Mediterranean coast around Tripoli, ultimate destinations are the developed economies of European Union (EU) and Britain, [28]. In recent years (2014-2020), one of the main transient routes for the migrants has been through Libya, proceeding toward resettlement countries. Pregnant women seek to reach Europe to provide citizenship for their new-born. Migrants seek work in Al Kufra to fund their onward migration, pooling funds to buy vehicles for the 1000 km trip to the Mediterranean.

**Figure 1.**
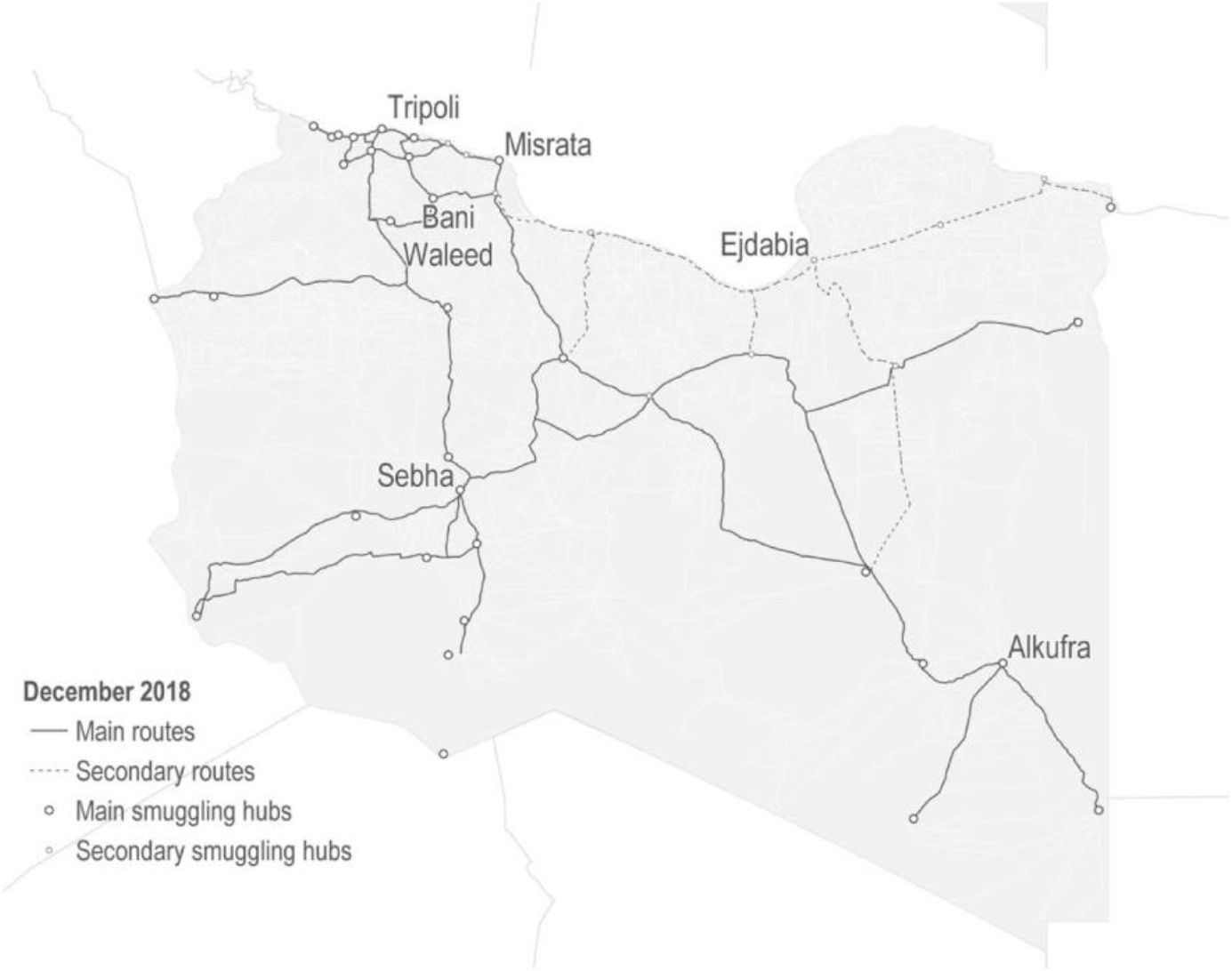
Example of migration routes from sub-Saharan Africa through Libya at the end of 2018, UNHCR, 2019 [30].

Local health officials recognize that the migrants arrive at Al Kufra in desperate condition, on foot, carrying only what has been essential for survival in crossing the desert. Infectious diseases are common including hepatitis, tuberculosis, STDs, and all manner of respiratory and enteric infections. In effort to control the spread of disease, migrants seeking documentation to permit work or for licenses for trade or commerce are required to obtain a health document indicating that they have presented at the local hospital for testing, the source of samples for this project. Documents must be presented at a checkpoint at the beginning of the road to the North. Despite this requirement, an unknown proportion, estimated ca. 2/3-3/4 of migrants, fearing travel restriction, do not present at the hospital and circumvent the checkpoint on foot to be picked up at a point beyond and out of sight of the checkpoint. The number of migrants, trying to reach Europe from Al Kufra, estimated from hospital records and checkpoint observations, are typically 500 migrants/day, with approximately 500/week remaining in Al Kufra, 100/month returning to countries of origin, resulting in approximately 2000/week passing the checkpoint. Currently increasing, the flow of African migrants into Al Kufra, city from neighboring countries, especially those arriving from intermediate and high endemic countries, is an additional burden on the local population in the host nations.

Migrants newly arrived from the neighboring and other sub-Saharan African countries have been cited as one of the major factors spreading the chronic liver disease and hepatocellular carcinoma worldwide and the risks of disease transmission to the local population [24, 10]. Collectively, migrants suffer from a variety of infectious diseases that are usually more prevalent in their own countries [12]. Migrants from Somali, and Eritrea in particular, have a high prevalence of viral hepatitis [15, 25, 29]. A previous study from Libya reported the prevalence and distribution of HCV genotypes among the North and sub-Saharan African migrants as they resided in the transit country before reaching a final destination country [10]. Infections with viral hepatitis, particularly hepatitis B (HBV) and hepatitis C (HCV), have become a universally established health challenge due to worldwide distribution, leading to similar types of chronic liver disease including hepatocellular carcinoma, particularly in economically poor countries [26].

The aim of this project was to determine HBV and HCV prevalence in newly arrived African migrants arriving at Al Kufra City from sub-Saharan regions where both viral infections are highly endemic.

## Materials and methods

This project was conducted for 12 months from January 01 to December 31,2019, at Al Kufra City hospital laboratory in a high migrant area of Libya. Migrants arriving at Al Kufra and seeking documentation, are required to attend the local hospital for a basic health examination supervised by the Libyan National Centre for Disease Control (CDC) following national ethical guidelines. With informed consent, a 5 mL blood sample is collected to identify infectious disease conditions. At sample collection socio-demographic characteristics (age, sex, and country of origin) are recorded. Following analysis for CDC requirements, serum samples are retained briefly before disposal. The sera were provided for unlinked anonymous testing (UAT) in this project and approved by the Office of the Faculty Undersecratary for Scientific Affairs, Bengazi University-Libyan, not requiring further ethical oversight. Retained sera were stored at -20° C until tested. In the 2019 calendar year serum samples from were analyzed from 3248 newly arrived migrants from different parts of sub-Saharan Africa.

The UAT serum samples were tested and confirmed for HBV and HCV. Screening for HBV used the “Advanced Quality™ One Step HBsAg Test” for detection of HBV surface antigen, an imunochromatograpic assay for serum or plasma samples (InTec Products Inc, Xiamen, Fujian P.R. China). Screening for HCV used the “Advanced Quality™ Rapid Anti-HCV Test” for detection of anti-HCV antibodies, a colloidal gold enhanced immunochromatographic assay for human serum or plasma (InTec Products Inc, Xiamen, Fujian P.R. China). Samples that were positive for HBV and HCV further tested using the “HBsAg ELISA Kit” and the “HCV Ab ELISA Kit, both are third-generation enzyme linked immunosorbent assays (ELISA), (BIONEOVAN Co., Ltd, Beijing, P.R. China). All screening positive samples were confirmed at least once to avoid false positive results and considered as positive only if the retest was positive. Positive and negative control serums provided with the assay kits for HBV and HCV tests were included in all assay runs following the manufacturer’s instructions to avoid false positive and negative results.

### Statistical Analysis

Data were analyzed using SPSS software version 25. Descriptive statistics were performed to describe the frequency of the different study variables, and to estimate the prevalence of HBV and HCV infections. The associations of the gender, age, and country of origin with hepatitis B surface antigen (HBsAg) and anti-hepatitis C virus (HCV) antibodies results as the dependent variable were tested using binary logistic regression. Odds ratio (OR) at 95% confidence interval (CI) was calculated and a p-value < 0.05 was considered significant.

### Ethical approval

As described above, written informed consent was obtained from study participants after briefing on CDC requirements for infectious disease screening. The nature of the UAT samples to be analyzed in this project were examined by the responsible ethics body of Benghazi Uinversity-Libyan. Recognizing the strict UAT control and unlinked demographic information with serum samples, the project was judged to not require further ethical oversight.

## Results

A total of 3248 newly arrived migrants from different parts of sub-Saharan Africa countries were included in the study, and among them 3135 were male, and 113 female 96.5% and 3.5%, respectively, and their age range was 18–53 years with a mean age of 33.1 ± 8.8 SD years. The population was divided by age into 3 groups, 18-28, 28-38, and 39-53, Table 1.

**Table 1.** Age and gender distribution of 3248 sub-Saharan African migrants newly arrived at Al Kufra, January to December 2019, travelling toward the Mediterranean and Europe.

| **Population Age Range** | **Male No. (%)** | **Female No. (%)** | **Total No. (%)** |
| --- | --- | --- | --- |
| 18-28 | 1054 | 78 | 1132 (34.9) |
| 29-38 | 1066 | 29 | 1095 (33.7) |
| 39-53 | 1015 | 6 | 1021 (31.4) |
| sub-Total | 3135 (96.5) | 113 (3.5) | 3248 |

The population was distributed in approximately equal thirds across the three age ranges, slightly skewed to the lower ages. Most of the migrants in this study were from central African countries (Chad and Sudan), 1901(58.5%). The west African countries (Burkina Faso, Niger, Mali, Nigeria, and Ghana) and east African countries (Eritrea, Somalia and Ethiopia) were the second largest migrant category, 890 (27.4%) and 457 (14.1%), respectively (Table 2). Over a period of 12 months, a total of 3248 newly arrived migrants from different parts of sub-Saharan Africa were screened for HBV and HCV. Out of these 761 were positive for HBsAg, and 1014 for anti-HCV rapid tests. The same prevalence was found by the ELISA confirming test. The percentage was calculated for both HBV and HCV positive subjects was 23.4% for HBV. and 31.2% for HCV.

**Table 2.** Countries of origin of 3248 sub-Saharan African migrants newly arrived at Al Kufra, January to December 2019, travelling toward the Mediterranean and Europe.

| **Region/Country** | **Total (%)** | **Male** | **Female** |
| --- | --- | --- | --- |
| **East Africa:** |  |  |  |
| Eritrea | 137 (4) | 108 | 29 |
| Somalia | 199 (6) | 174 | 25 |
| Ethiopia | 121 (3.7) | 112 | 9 |
| **Central Africa:** |  |  |  |
| Chad | 1032 (32) | 1009 | 23 |
| Sudan | 869 (27) | 848 | 21 |
| **West Africa:** |  |  |  |
| Burkina Faso | 173 (5) | 172 | 1 |
| Niger | 450 (14) | 446 | 4 |
| Mali | 182 (5.6) | 181 | 1 |
| Nigeria | 64 (2) | 64 | 0 |
| Ghana | 21 (1) | 21 | 0 |
| **Total** | 3248 (100) | 3135 | 113 |

### HBV Prevalence

The overall prevalence of HBV in the 3248 migrant population was 23.4%. The HBV prevalence by HBsAg positivity was higher among male participants, 23.9% (750/3135) than among female participants, 9.7% (11/113), (Table 3). The difference was significant (P = 0.001) with odds of males being HBV positive nearly twice in comparison to females (Table 3). In the present study, statistically significant differences were observed in the age group of 18–28 [25.35% (287/1132)] and [31.6% (346/1095)] 29–38-years old participants with HBV infection (OR = 1.57, CI 95% = 1.23-2.03, P = 0.001 and OR = 2.176, CI95% = 1.70-2.78, P = 0.001) compared to age group 39+ [12.54% (128/1021)], respectively. In east African migrants, the prevalence of HBV was 55.57% (254/457). The prevalence of HBV in west African and in central African migrants was 35.6% (317/890) and 9.99% (190/1901), respectively.

**Table 3.** Seroprevalence of hepatitis B surface antigen (HBsAg) among African migrants transiting Al Kufra, Libya, by demographic characteristics and regions of origin.

| Category | Prevalence of HBsAg<br>No (%) |  | OR | 95%CI |  | P<br>value |
| --- | --- | --- | --- | --- | --- | --- |
|  | positive<br>761 (23.4) | negative<br>2487 (76.6) |  | Lower | Upper |  |
| **Age (yrs):** |  |  |  |  |  |  |
| 39+ | 128 (16.8) | 893 (35.9) | 1.00 |  |  |  |
| 29–38 | 346 (45.5) | 749 (30.1) | 2.17 | 1.70 | 2.78 | .001 |
| < =28 | 287 (37.7) | 845 (34.0) | 1.58 | 1.23 | 2.03 | .001 |
| **Gender:** |  |  |  |  |  |  |
| Females | 11 (1.4) | 102 (3.9) | 1.00 |  |  |  |
| Males | 750 (98.6) | 2385 (96.1) | 3.39 | 1.83 | 6.24 | .001 |
| **Region:** |  |  |  |  |  |  |
| (CA) | 190 (24.9) | 1711 (68.8) | 1.00 |  |  |  |
| (WA) | 317 (41.7) | 573 (23.0) | 4.18 | 3.39 | 5.16 | .001 |
| (EA) | 254 (33.4) | 203 (8.2) | 10.76 | 8.42 | 13.75 | .001 |

Central Africa (CA), West Africa (WA), East Africa (EA). Confidence Interval (CI), Odds Ratio (OR), Hepatitis B surface antigen (HBsAg)

### HCV Prevalence

The overall prevalence of HCV in the 3248 migrant population was 31.2 (Table 3). The prevalence of HCV in migrants from west African countries was 53.82% (479/890). The prevalence of anti-HCV positivity (30.8%) among male participants was lower than female participants (41.6%) (Table 4), and the difference was significant (OR = .54, CI 95% = 0.35-.84, P = 0.006). Statistically, significant association could be seen in the age group of 18–28 [31% (351/1132)] and [47.21% (517/1095)] 29–38-years old participants with HCV infection (OR = 2.10, CI 95% = 1.65-2.67, P = 0.001 and OR = 3.22, CI95% = 2.55-4.07, P = 0.001) compared to other age groups, respectively.

**Table 4.**
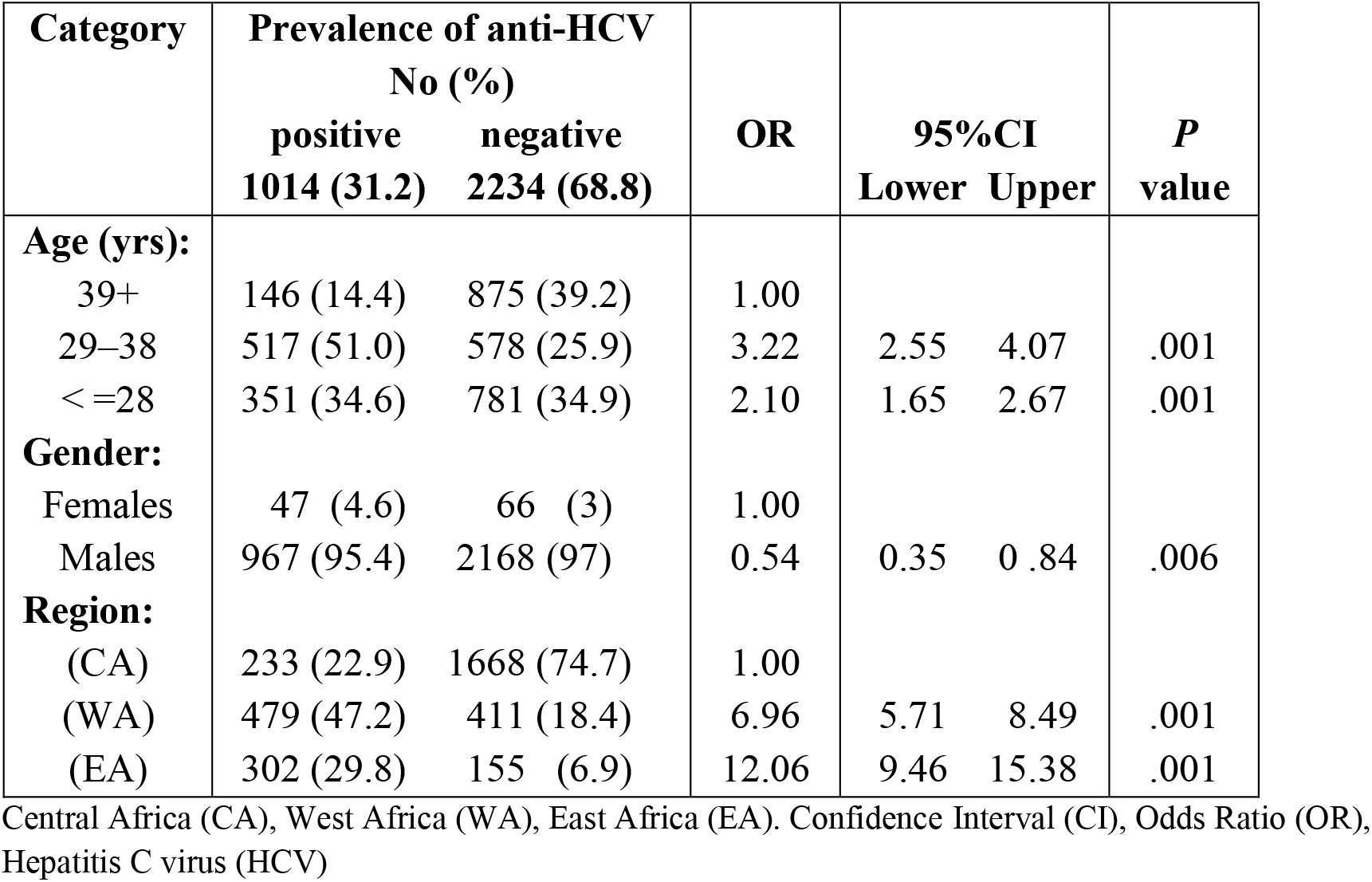
Seroprevalence of anti-HCV antibodies among African migrants transiting Al Kufra, Libya, by demographic characteristics and regions of origin.

| **Category** | **Prevalence of anti-HCV**<br>**No (%)** |  | **OR** | **95%CI** |  | **P**<br>**value** |
| --- | --- | --- | --- | --- | --- | --- |
|  | **positive**<br>**1014 (31.2)** | **negative**<br>**2234 (68.8)** |  | **Lower** | **Upper** |  |
| **Age (yrs):** |  |  |  |  |  |  |
| 39+ | 146 (14.4) | 875 (39.2) | 1.00 |  |  |  |
| 29–38 | 517 (51.0) | 578 (25.9) | 3.22 | 2.55 | 4.07 | .001 |
| < =28 | 351 (34.6) | 781 (34.9) | 2.10 | 1.65 | 2.67 | .001 |
| **Gender:** |  |  |  |  |  |  |
| Females | 47 (4.6) | 66 (3) | 1.00 |  |  |  |
| Males | 967 (95.4) | 2168 (97) | 0.54 | 0.35 | 0.84 | .006 |
| **Region:** |  |  |  |  |  |  |
| (CA) | 233 (22.9) | 1668 (74.7) | 1.00 |  |  |  |
| (WA) | 479 (47.2) | 411 (18.4) | 6.96 | 5.71 | 8.49 | .001 |
| (EA) | 302 (29.8) | 155 (6.9) | 12.06 | 9.46 | 15.38 | .001 |

Central Africa (CA), West Africa (WA), East Africa (EA). Confidence Interval (CI), Odds Ratio (OR), Hepatitis C virus (HCV)

The east African migrants showed the highest prevalence rates of HCV 66.08% (302/457), while the central African migrants showed the lowest prevalence rates 12.26% (233/1901). According to binary logistic regression analysis, relative to the reference category of central Africa, statistically significant differences were observed for HBV and HCV infections in terms of geographical regions, HCV and HBV prevalence were in higher in migrants from different parts of east African countries than migrants from central African countries (OR = 12.06, CI 95% = 9.46-15.38, P = 0.001 and OR = 10.76, CI 95% = 8.42-13.75, P = 0.001). Furthermore, HCV and HBV prevalence were higher in the west African migrant population than migrants from different parts of central African countries (OR = 6.96, CI 95% = 5.71-8.49, P = 0.001 and OR = 4.18, CI 95% = 3.39-5.16, P = 0.001) (Table 2).

## Discussion

Overall, the basic features of hepatitis B and C, the diseases that they cause, the consequences of infection, and their epidemiology…distribution, prevalence by region, and risk factors…are well-established [7, 8, 9, 26]. Epidemiology of both HBV and HCV are characterized by a wide range of prevalence related closely to economic, social, and sanitation conditions regionally. Prevalence of HBV globally ranges from < 2% (e.g., 0.2% in the USA, 1% avg in Europe) to 8% in areas of Asia and Africa. Prevalence of HCV globally ranges from < 1% (e.g., Northern Europe, Middle East, India) to >5% (e.g., areas of Africa, central Asia, Tibet). Libya overall, where this work was conducted, has “Low Intermediate” HBV prevalence, 1.5-2%, and “Low Moderate” HCV prevalence, also 1.5-2%. Against the background levels of HBV and HBC prevalence in the Libyan at large, the data presented here show that in 3248 migrants in the 2019 calendar year prevalence was much higher with nearly 1/4 (23.4%) having evidence of HBV infection and nearly 1/3 (31.2%) having evidence of HCV infection.

Important questions in light of these data are: 1) are the data representative of migrants? 2) are the data representative of the population in regions of origin? 3) what is the significance for destination populations? and 4) what actions/policies do the data suggest?

1. The data are from a large population, 3248 samples analyzed over the calendar year. However, based on the estimate of arrivals at Al Kufra of ca. 500/day, the samples analyzed are a small fraction of the total population i.e., about 10/day. This represents the population presenting at the local hospital to obtain documentation, ultimately needed to pass the departure checkpoint. The population passing the checkpoint estimated at ca. 2000/month (ca. 100/day) is thus 1/4 of the total migrants passing Al Kufra. Whether the prevalence of 23% HBV and 31% HCV are representative is a matter of speculation.
2. According to published reports HBV and HCV prevalence in sub-Saharan populations varies widely depending on the specific population sampled. It appears that although both HBV and HCV rates among the general populations in countries of origin are in the range of 2 to 10% for HBV and 2 to 15% for HCV, infection rates in the migrant populations are more reflective of higher risk groups. Local observers describe the general condition of arriving migrants specifically at Al Kufra as depleted and sharing everything including clothing, eating, and personal care items offering ample opportunity for blood born exposure.
3. The significance of high HBV and HCV prevalence in migrant populations to destination country residents has been examined by European health agencies concluding that indeed, due to the high prevalence, migrant infections increase the prevalence in the overall destination countries [11]. However, the risk of transmission is highest among immigrant groups and not significantly to resident populations. Overall impact is country specific related to the immigrant proportions in destination countries and to the HBV and HCV prevalence in the native populations.
4. Actions/Policies. The effectiveness of effort allocated to public health related to HBV and HCV depends on the accuracy of modelling supporting policy and resulting actions. In turn, the accuracy of modelling is controlled by input data. The ability to understand the significance of HBV and HCV introduced to destination country populations depends on understanding HBV and HCV prevalence in migrants [11]. The population studied here appears relatively unbiased, early at a collection point in migration before dispersal, and provides a unique view of migrants from the areas described.

This study shows significant rates of HCV and HBV prevalence in African migrants gathered in Libya from many sub-Saharan African countries, an overall estimated HBV and HCV prevalence of 23.4% and 31.2% respectively, with higher prevalence in migrants from the East African region (HCV,66.1%, and HBV, 55.6%), and in the West African region (HCV, 53.8% and HBV, 35.6%) and lower rates in the central African region (HCV, 12.3% and HBV, 10.0%). The regional hepatitis B surface antigen (HBsAg) and HCV-antibody estimates in our study were higher than those reported in many sub-Saharan countries of Africa. Suggesting that the migrants included in our study, ones sufficiently desperate to risk the journey, have differed from the populations at large in their countries of origin.

## Conclusion

Migrants originating in sub-Saharan Africa are of great interest because the region is reported to have the high HBV and HCV prevalence rate. This study shows clearly that HBV and HCV prevalence in migrants significantly exceed even the relatively high viral hepatitis prevalence sub-Saharan African countries. We measured the overall HBV prevalence of 23.4% and HCV prevalence of 31.2% in a large and representative population of migrants. The findings of this study suggest that migrants from high HBV and HCV prevalence countries represent an important risk group for HCV and HBV infection as they proceed toward and integrate into European destination populations.

## Data Availability

All data produced in the present work are contained in the manuscript

## Acknowledgment

The contribution of Fayizah A. M. Alkaddouh in assisting implementation of essential project activities is gratefully acknowledged.

## Notes

### Competing Interest Statement

The authors have declared no competing interest.

### Funding Statement

This study did not receive any funding

### Author Declarations

A waiver of ethical approval was received from the Office of the Faculty Undersecratary for Scientific Affairs, Bengazi University-Libyan.

## References

1. Abdulaziz Q. Alodini. Prevalence of Hepatitis B Virus (HBV) and Hepatitis C Virus (HCV) Infections among Blood Donors at Al-Thawra Hospital Sana’a City-Yemen. Yemeni Journal for Medical Sciences (2012).

2. Ahmed S, Sanyal R, Scedrou P, Ahmed M. Hepatitis B surface antigen in South Sudan. J Commun Disord. 1984;16(4):330–1.

3. Ali A, Ahmad H, Ali I, Khan S, Zaidi G, Idrees M. Prevalence of active Hepatitis c virus infection in district Mansehra Pakistan. Virol J 2010;7:334.

4. Blumberg BS. Hepatitis B - The hunt for a killer virus. Princeton: Princeton University Press; 2002.

5. Burnett R, Francois G, Kew M, Leroux-Roels G, Meheus A, Hoosen A, et al. Hepatitis B virus and human immunodeficiency virus co-infection in sub-Saharan Africa: a call for further investigation. Liver Int. 2005;25(2):201–13.

6. CDC, 2021. Centers for Disease Control and Prevention (CDC). Global Viral Hepatitis: Millions of People are Affected. https://www.cdc.gov/hepatitis/global/index.htm#ref01 (Accessed 7 December 2021)

7. CDC. Epidemiology and prevention of vaccine-preventable diseases (The pink book). 10th edition. Waldorf (MD): Public Health Foundation; 2008.

8. CDC. Yellow Book Chapter 4-Hepatitis B, 2020: Health Information for International Travel. New York: Oxford University Press; 2017.

9. CDC. Yellow Book Chapter 4-Hepatitis c, 2020: Health Information for International Travel. New York: Oxford University Press; 2017.

10. Daw MA, et al. Hepatitis C Virus in North Africa: An Emerging Threat. The Scientific World Journal. Volume 2016, Article ID 7370524, 11 pages. http://dx.doi.org/10.1155/2016/7370524.

11. ECDC. Epidemiological assessment of hepatitis B & C among migrants in the EU/EEA. Stockholm: ECDC; 2016. https://www.ecdc.europa.eu/sites/default/files/media/en/publications/Publications/epidemiological-assessment-hepatitis-B-and-C-among-migrants-EU-EEA.pdf (Accessed Jan 10, 2021).

12. Gushulak BD, MacPherson DW. Globalization of infectious diseases: the impact of migration. Clin Infect Dis 2004 Jun 15; 38(12):1742e8. PMID: 15227621.

13. Hajarizadeh B, Grebely J, Dore GJ. Epidemiology and natural history of HCV infection. Nat Rev Gastroenterol Hepatol 2013; 10(9): 553–62.

14. Houghton M. Discovery of the hepatitis C virus. Liver Int. 2009 Jan;29 Suppl 1:82–8. doi: 10.1111/j.1478-3231.2008.01925.x.

15. Howell J, Ladep NG, Lemoin M, Thursz MR, Taylor-Robinson SD. Hepatitis B in sub-Saharan Africa. S Sudan Med J. 2014;7(3):59–61.

16. Jamma S, Hussain G, Lau DT. Current Concepts of HBV/HCV Coinfection: Coexistence, but Not Necessarily in Harmony. Current Hepatitis Reports. 2010;9(4):260–269.

17. Junejo SA, Khan NA, Lodhi AA. Prevalence of Hepatitis B and C infection in patients admitted at Tertiary Eye Care Centre: A hospital-based study. Pak J Med Sci 2009;25(4):597–600.

18. Kefene H, Rapicetta M, Rossi G, Bisanti L, Bekura D, Morace G, et al. Ethiopian national hepatitis B study. J Med Virol. 1988;24(1):75–84.

19. Mukherjee S and Dhavan V. Hepatitis C. eMed J 2004. Available from URL: http://www.emedicine.com/gastroentrology/hepatitisC.

20. Owiti JA, et al. Illness perceptions and explanatory models of viral hepatitis B & C among migrants and refugees: a narrative systematic review. BMC Public Health 2015) 15:151

21. Riou J, Aït Ahmed M, Blake A, Vozlinsky S, Brichler S, Eholié S, et al. Hepatitis C virus seroprevalence in adults in Africa: a systematic review and meta-analysis. J Viral Hepat 2015 Oct 19. doi:10.1111/jvh.12481. PMID:26558905. doi: 10.1371/journal.pone.0141715.

22. Saaed FMA, Ongerth JE. Giardia and Cryptosporidium in children with diarrhea, Al Kufra, Libya, an agricultural North African city; Volume 222, Issue 5, June 2019, Pages 840–846.

23. Saleh MG et al. High prevalence of hepatitis C virus in the normal Libyan population. Transactions of the Royal Society of Tropical Medicine and Hygiene, 1994, 88:292–294.

24. Seeger C, Mason WS. Hepatitis B virus Biology. Micro Mol Biol Rev 2000; 64: 51–68.

25. Sharma S, Carballo M, Feld JJ, Janssen HL. Immigration and viral hepatitis. J Hepatol. 2015;63(2):515–22.

26. Te, H. S., and D. M. Jensen. “Epidemiology of hepatitis B and C viruses: a global overview,” Clinics in Liver Disease, vol. 14, no. 1, pp. 1–21, 2010.

27. Tong CY, Khan R, Beeching NJ, Tariq WU, Hart CA, Ahmad N, et al. The occurrence of hepatitis B virus and HCV in Pakistani patients with chronic live disease and hepatocellular carcinoma. Epidemiol Infect. 117:327–32, 1996

28. Torelli, SM, 2018. Migration through the Mediterranean: Mapping the EU response. European Council on Foreign Relations (ECFR.eu), April 2018. https://ecfr.eu/special/mapping_migration/# (Accessed, Jan 2, 2022).

29. UNHCR. Ethiopia refugee response plan; 2018. p. 1–68.

30. UNHCR, 2018. Mixed Migration Routes and Dynamics in Libya in 2018. United Nations, High Commission on Refugees, 2019. Available at: https://reliefweb.int/sites/reliefweb.int/files/resources/impact_lby_report_mixed_migration_routes_and_dynamics_in_2018_june_2019.pdf. (Accessed Jan 11, 2022).

31. WHO: Global surveillance and control of hepatitis C. Report of a WHO Consultation organized in collaboration with the Viral Hepatitis Prevention Board, Antwerp, Belgium. J Viral Hepat 1999, 6(1):35–47.

32. World Health Organization. (2021). Global progress report on HIV, viral hepatitis and sexually transmitted infections, 2021: accountability for the global health sector strategies 2016–2021: actions for impact: web annex 1: key data at a glance. World Health Organization. https://apps.who.int/iris/handle/10665/342808. (Accessed, Jan. 10, 2022)

